# Natural Language Processing for Mimicking Clinical Trial Recruitment in Critical Care: A Semi-automated Simulation Based on the LeoPARDS Trial

**DOI:** 10.1101/19005603

**Authors:** Hegler Tissot, Anoop Shah, Ruth Agbakoba, Amos Folarin, Luis Romao, David Brealey, Steve Harris, Lukasz Roguski, Richard Dobson, Folkert Asselbergs

## Abstract

Clinical trials often fail on recruiting an adequate number of appropriate patients. Identifying eligible trial participants is a resource-intensive task when relying on manual review of clinical notes, particularly in critical care settings where the time window is short. Automated review of electronic health records has been explored as a way of identifying trial participants, but much of the information is in unstructured free text rather than a computable form. We developed an electronic health record pipeline that combines structured electronic health record data with free text in order to simulate recruitment into the LeoPARDS trial. We applied an algorithm to identify eligible patients using a moving 1-hour time window, and compared the set of patients identified by our approach with those actually screened and recruited for the trial. We manually reviewed clinical records for a random sample of additional patients identified by the algorithm but not identified for screening in the original trial. Our approach identified 308 patients, of whom 208 were screened in the actual trial. We identified all 40 patients with CCHIC data available who were actually recruited to LeoPARDS in our centre. The algorithm identified 96 patients on the same day as manual screening and 62 patients one or two days earlier. Analysis of electronic health records incorporating natural language processing tools could effectively replicate recruitment in a critical care trial, and identify some eligible patients at an earlier stage. If implemented in real-time this could improve the efficiency of clinical trial recruitment.

## 1 Introduction

Randomised clinical trials can provide robust evidence of the effectiveness of medicines and other treatments, but are expensive to conduct and may fail to recruit a sufficient number of appropriate patients to have adequate statistical power. Clinical trials units try to use a variety of techniques to increase patient recruitment, such as increasing the awareness amongst patients and clinicians [1]. However, identification of suitable patients can be resource-intensive, often relying on manual review of clinical notes to identify potentially eligible patients, where the information may be split over different systems. This can be particularly difficult in emergency and critical care settings, when it is important to identify eligible patients early so that the window of opportunity is not missed [2]. Staff shortages and inconvenient timing can potentially lead to eligible participants being missed [3].

Electronic heath records (EHRs) are increasingly used for research [4] and have been proposed as a way of improving trial recruitment, either via a patient-centric approach or in the form of decision support for clinicians, such as point-of-care alerts [5]. Algorithms to identify trial participants may reduce the human resource needed for identifying patients earlier. Patient characteristics extracted from EHR databases can be mapped to trial information derived from study eligibility criteria [6, 7]. However, much of the information in EHRs is unstructured, in the form of free text, rather than in a structured form. Natural language processing (NLP) techniques can extract relevant information from free text, but cannot be relied upon to be completely accurate because of typographical errors and nuances of human language. However, NLP may be used within algorithms to pre-screen potential trial participants, reducing the number of patient records that need manual review [8–10].

In this study we developed an NLP pipeline and patient selection algorithm to simulate screening and recruitment for the LeoPARDS trial [11], a trial of an intervention for life-threatening infections (sepsis). We chose this trial as the exemplar because it required heterogenous clinical data to be interpreted within narrow time windows. We aimed to test whether NLP in combination with electronic structured data could assist in trial recruitment in critical care. The simulation was conducted within one of the LeoPARDS trial sites, University College London Hospitals NHS Foundation Trust (UCLH). UCLH is is a teaching hospital and part of a National Institute for Health Research (NIHR) Biomedical Research Centre (BRC), and is leading a collaboration across multiple BRCs to curate a critical care research database within the NIHR Health Informatics Collaboration (CCHIC) [12].

## 2 Methods

### 2.1 Data Sources and Informatics Infrastructure

The Critical Care Health Informatics Collaboration (CCHIC) is a research platform comprising EHR patient data from critical care units at five large BRCs (Cambridge, Guys/Kings/St Thomas’, Imperial, Oxford, and UCLH) [12]. Data is available from 2014 onwards, and is extracted in a standardised format, curated into a research-ready database and provided to researchers under an ethical and governance framework to for observational research. The CCHIC database has been approved by a Research Ethics Committee. The CCHIC dataset includes 108 hospital, unit, patient and episode descriptors (recorded once per admission) and 154 time-varying variables including physiological measurements, laboratory tests, nursing activities and drug administration.

For this study we used structured UCLH EHR from CCHIC and unstructured free text from the UCLH critical care EHR (the IntelliVue Clinical Information Portfolio (ICIP) by Phillips). We used free text recorded in the following parts of the EHR: problem lists, event timeline, reason for admission, admission history, past medical history, and pre-admission medication.

Stuctured and free text data from the EHR were combined into a searchable indexed repository using the CogStack [13] platform, which contains pipelines for document processing and indexing, fast text searching, and distributed analysis. We used the SemEHR [14] biomedical document processing system on CogStack, with Elasticsearch^1^ for full free text search to explore text and annotations and Bio-Yodie [15] (an NLP application) to annotate text using the Unified Medical Language System (UMLS) [16]. SemEHR contextualises each mention of a UMLS concept with the experiencer (patient or other), affirmation status (affirmed, negative or hypothetical) and temporality (past or recent). We only used affirmative (non negative) UMLS concepts that were experienced by the patient in this study.

We developed an application using CogStack to mimic the trial screening process. An ontological view of all the contextualized concepts was used to perform patient eligibility searches, based on a matching algorithm for selection criteria. We compared potentially eligible patients identified by CogStack with those included in the original LeoPARDS trial, during the intersection of time periods between trial recruitment and the CCHIC data (June 2014 to December 2015). All analysis on the EHR data was carried out by researchers blinded to the trial recruitment log, with no involvement in the original trial.

### 2.2 The LeoPARDS Trial

The LeoPARDS trial (Levosimendan for the Prevention of Acute oRgan Dysfunction in Sepsis) investigated whether a 24-hour infusion of levosimendan improved organ dysfunction in septic shock [11]. The trial screened 2,382 patients in 2014–2015 across 31 centres and recruited 526, with 47 patients recruited from UCLH. The primary outcome was the mean daily Sequential Organ Failure Assessment (SOFA) score, which is used to track the evolution of organ dysfunction, and the study showed no significant difference between the levosimendan group and the placebo group (mean difference in SOFA score, 0.61; 95%CI, −0.07 to 1.29).

Recruitment into the LeoPARDS study required the identification of patients with new onset septic shock within 24 hours, so that they could be randomised to the study drug or placebo. Eligible patients were identified by dedicated research nurses who reviewed the notes of all new admissions to the critical care unit, which took 4–6 hours per day. The selection criteria are shown in Table 1. The *Inclusion Criteria* targeted adult patients (`≥`18 years) who required vasopressor support for the management of sepsis despite fluid resuscitation, using a previously accepted definition of septic shock [17]. The *Exclusion Criteria* were specified in order to exclude patients in whom the trial therapy was inappropriate or potentially dangerous, or if their condition might make the outcome of the trial more difficult to interpret.

**Table 1:** Selection criteria for the LeoPARDS trial [11].

| Inclusion Criteria | Exclusion Criteria |
| --- | --- |
| (A) Fulfil 2/4 of the criteria of the systemic inflammatory response syndrome (SIRS) due to known or suspected infection within the previous 24 hours. The SIRS criteria are:<br>1. fever ($> 38^{\circ}\text{C}$) or hypothermia ($< 36^{\circ}\text{C}$),<br>2. tachycardia (heart rate $> 90$ beats per minute),<br>3. tachypnoea (respiratory rate $> 20$ breaths per minute or $\text{PaCO}_2 < 4.3$ kPa) or need for mechanical ventilation, and<br>4. abnormal leukocyte count ($> 12,000$ cells/$\text{mm}^3$, $< 4000$ cells/$\text{mm}^3$, or $> 10\%$ immature [band] forms).<br>(B) Hypotension, despite adequate intravenous fluid resuscitation, requiring treatment with a vasopressor infusion (e.g. noradrenaline / adrenaline / vasopressin analogue) for at least four hours and still having an ongoing vasopressor requirement at the time of randomisation. | (A) more than 24 hours since meeting all the inclusion criteria;<br>(B) end-stage renal failure at presentation (previously dialysis-dependent);<br>(C) severe chronic hepatic impairment (Child-Pugh class C);<br>(D) a history of Torsades de Pointes;<br>(E) known significant mechanical obstructions affecting ventricular filling or out-flow or both;<br>(F) treatment limitation decision in place (e.g. Do Not Resuscitate or not for ventilation/dialysis);<br>(G) known or estimated weight $> 135$ kg;<br>(H) known to be pregnant;<br>(I) previous treatment with levosimendan within 30 days;<br>(J) known hypersensitivity to levosimendan or any of the excipients;<br>(K) known to have received another investigational medicinal product within 30 days or currently in another interventional trial that might interact with the study drug – potential co-enrolment into other studies would be considered on an individual study basis. |

### 2.3 Patient Eligibility

Patients were eligible for LeoPARDS if they fulfilled at least two of the four criteria of the systemic inflammatory response syndrome (SIRS) due to known or suspected infection within the previous 24 hours, and had hypotension, persisting despite adequate intravenous fluid resuscitation, requiring vasopressor treatment. SIRS is defined by structured physiological or biomarker data including high heart rate (tachycardia), high respiratory rate (tachypnoea), or requirement for ventilation, fever or hypothermia, and high white cell count.

We operationalised ‘known or suspected infection’ as a recent diagnosis of infection from SemEHR (with specific types and sites of infection listed as search terms), and administration of a vasopressor for at least four hours during the previous 24 hours, and ongoing at the time of criteria evaluation (assuming that all patients on vasopressors would already have received adequate fluid resuscitation). Patients were evaluated for eligibility every hour from the start of their ICU admission. As long as the first matching regarding the selection criteria for each patient was triggered, the patient was included in the automatic screening.

We then applied the LeoPARDS exclusion criteria using structured and unstructured data as follows: end stage renal failure, dialysis, torsades de pointes, mitral stenosis, aortic stenosis or severe hepatic impairment (using either *recent* or *past* temporal context provided by SemEHR), or pregnancy (using only *recent* temporal context). We additionally identified patients with severe hepatic impairment by the presence of any two of bilirubin `≥` 34.2 micromol/L (CCHIC structured data), ascites or encephalopathy. This is an approximation of Child-Pugh class C, assuming encephalopathy is severe and ascites is moderate, and the international normalised ratio and albumin are in the middle of the scoring ranges. However, some exclusion criteria (items I, J, and K from Table 1) were not taken into account, as we were unable to find any related UMLS clinical concepts within the available ICU clinical notes.

### 2.4 Timeline Simulation

UCLH was involved in recruiting patients for the LeoPARDS between June 2014 and December 2015. Eligibility was temporally constrained to patients with new onset septic shock identified within 24 hours. The actual recruitment process was time-consuming, and in theory required each patient to be assessed for eligibility every hour – the same frequency in which some vital sign measures are collected. In our study, we applied a sliding a 24-hour window in the algorithm that simulated the reviewing process for each patient in the critical care unit (see pseudocode in Fig. 1). Dates and periods of times were described using the TimeML^2^ notation [18].

**Figure 1:**
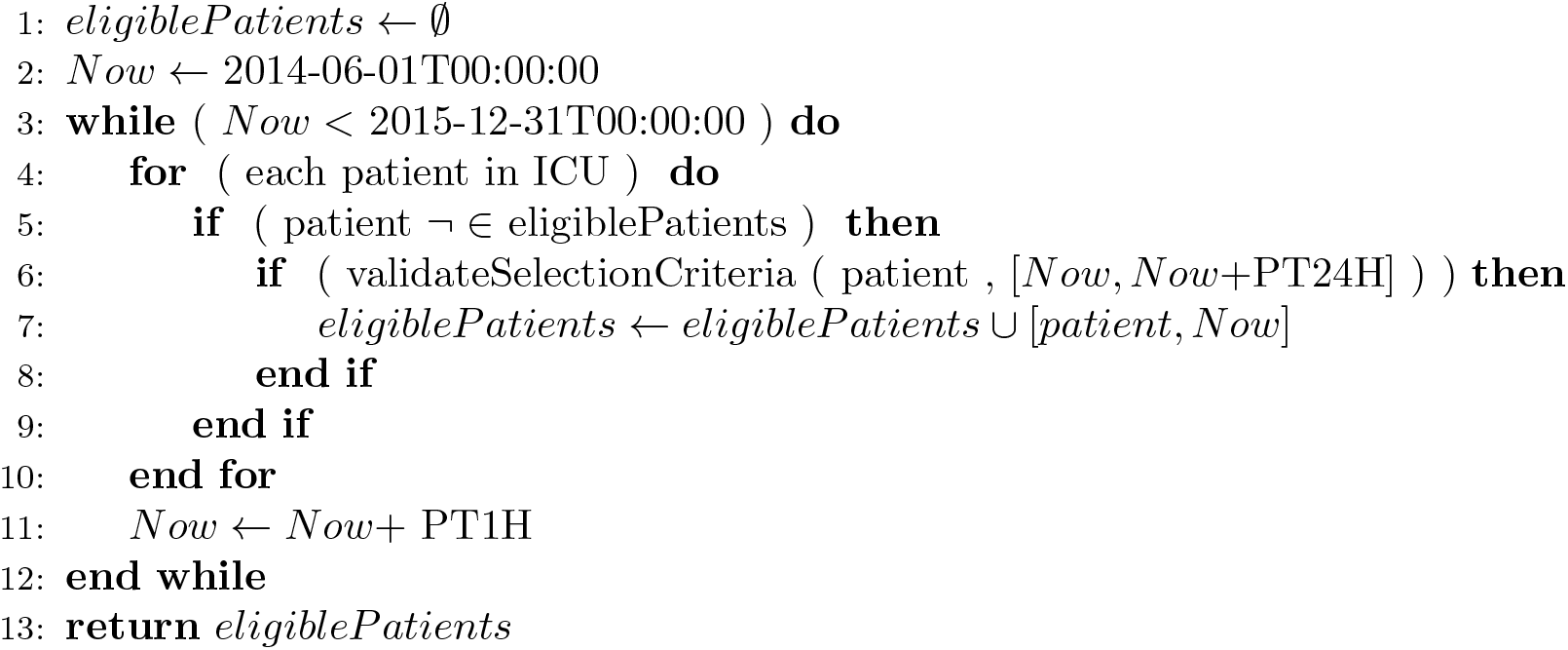
Sliding 24-hour-window for screening simulation pseudocode. Dates and periods of times are described using TimeML notation (e.g. `PT1H` means a time period of 1 hour duration, `P1D` is a 1 day time period, and `PT24H` is a 24 hour time period).

Each patient in the critical care unit (line 4) that has not been already included in the eligibility list (condition in line 5) was validated against the selection criteria for the current 24-hour sliding window (line 6). If the patient satisfied the inclusion criteria, he/she was included in the eligibility list with the corresponding earliest reference date for recruitment (variable *Now* in line 7). Exclusion criteria flags were returned as part of the eligibility conditions in order to be manually verified.

### 2.5 Technical Implementation of Eligibility Criteria

Our approach was designed to support an unlimited recursive nested set of conditional clauses connected by grouping logical operators. Partial matches and temporal constraints were also required as part of the formal criteria specification.

The selection criteria for the LeoPARDS trial was designed by following an inner hierarchical structure of conditional components. The two main components describe the set of conditions for each of the inclusion/exclusion conditions. The inclusion criteria comprise the default mandatory component for defining patient eligibility, requiring at least an inner logical group or an inner logical specification. The exclusion criteria are used as a complementary component comprising an inner logical group of conditions, specifying the set of patients to be subtracted (or flagged) from the initial cohort matching the inclusion conditions.

In order to formally describe the inclusion and exclusion criteria for the LeoPARDS trial, we designed a set of logical compounding functions (LCF). LCFs group a set of logical conditions that are individually evaluated and logically combined in order to determine whether the given criteria (logical set) result is *True* or *False*. Each LCF works as a grouping logical operator over a set of logical conditions, and each LCF result can be hierarchically combined to design more complex logical operations. The proposed LCFs are described below – *n* is a numerical constraint parameter and *L* is the set of logical conditions to be evaluated (all LCFs return *False* when *L* = ∅):

- `MIN`(*n, L*): each logical condition *c* ∈ *L* is logically evaluated, resulting *True* when at least *n* conditions from *L* result *True*;
- `MAX`(*n, L*): each logical condition *c* ∈*L* is logically evaluated, resulting *True* when no more than *n* conditions from *L* result *True*;
- `ALL`(*L*): results *True* if, and only if, there is no condition *c* ∈ *L* logically evaluated resulting *False*;
- `ANY`(*L*): results *True* when there is at least one logical condition *c* ∈ *L* that is logically evaluated resulting *True* – equivalent to: *MIN* (1, *L*);
- `ONE`(*L*): results *True* if there is only one condition *c* ∈ *L* that is logically evaluated resulting *True*, all the other conditions resulting *False* – equivalent to: *MAX*(1, *L*);
- `NOT`(*L*): results *True* if, and only if, there is no condition *c* ∈ *L* logically evaluated resulting *True* – equivalent to: *MAX*(0, *L*).

In addition to the LCFs described above, we defined a textual contextualised search function that looks for specific resulting UMLS concepts from SemEHR:

- `CONTEXT`(*umls, temporality*): is a textual contextualised search condition that matches annotated documents (free text notes) against one or more UMLS concepts (*umls* parameter) in a given time constraint (*temporality* parameter) – *temporality* can be set as *past* or *recent*, from which *recent* takes into account any UMLS concepts mentioned in any documents dated up to the last 72 hours from the reference screening date (variable *Now* in Fig. 1) set as “recent” by SemEHR, whereas *past* considers any historical occurrences of the given UMLS concepts. When *temporality* is not given, *Context* searches for any mention of the given UMLS identifiers that have been experienced by the patient.

We started by using the proposed LCFs to design the primary filters required to match patients according to the inclusion criteria. Primary conditions are supported by structured data points available in CCHIC. Table 2 formally describes each filter in terms of logical conditions coupling variables, logical operators, and grouping LCFs.

**Table 2:**
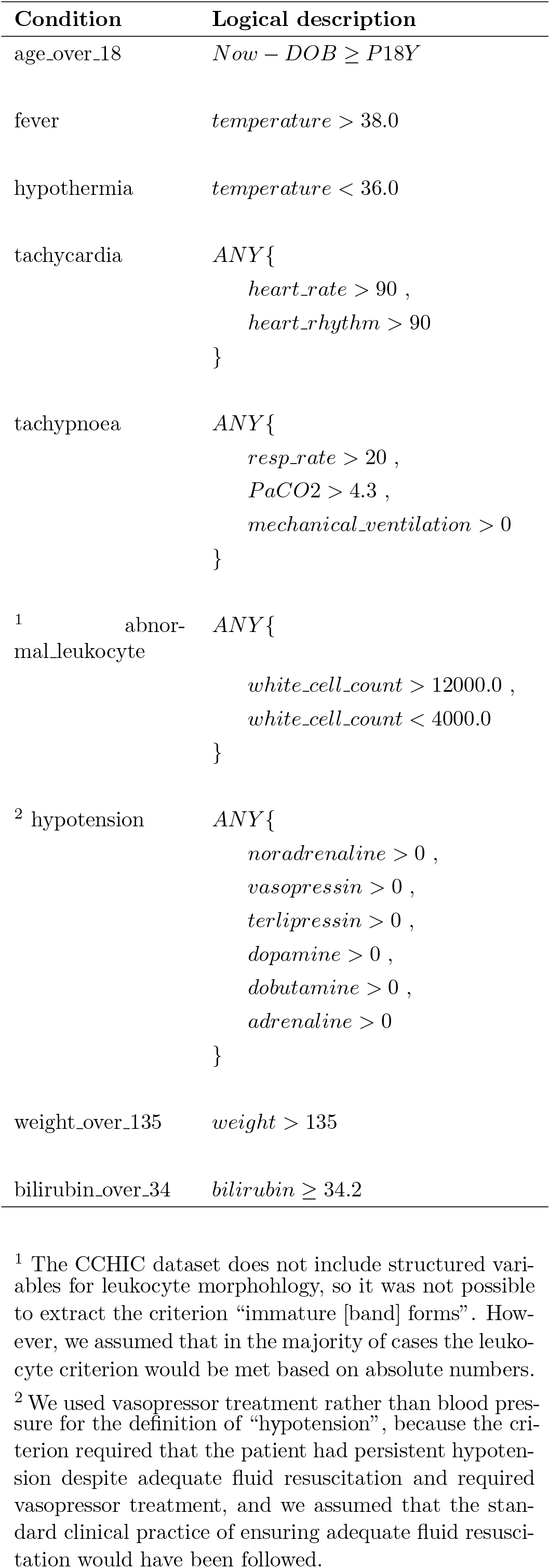
Logical conditions used to design the eligibility criteria.

| Condition | Logical description |
| --- | --- |
| age_over_18 | $Now - DOB \geq P18Y$ |
| fever | $temperature > 38.0$ |
| hypothermia | $temperature < 36.0$ |
| tachycardia | $ANY\{$<br>$heart\_rate > 90 ,$<br>$heart\_rhythm > 90$<br>$\}$ |
| tachypnoea | $ANY\{$<br>$resp\_rate > 20 ,$<br>$PaCO2 > 4.3 ,$<br>$mechanical\_ventilation > 0$<br>$\}$ |
| <sup>1</sup> abnormal_leukocyte | $ANY\{$<br>$white\_cell\_count > 12000.0 ,$<br>$white\_cell\_count < 4000.0$<br>$\}$ |
| <sup>2</sup> hypotension | $ANY\{$<br>$noradrenaline > 0 ,$<br>$vasopressin > 0 ,$<br>$terlipressin > 0 ,$<br>$dopamine > 0 ,$<br>$dobutamine > 0 ,$<br>$adrenaline > 0$<br>$\}$ |
| weight_over_135 | $weight > 135$ |
| bilirubin_over_34 | $bilirubin \geq 34.2$ |

<sup>1</sup> The CCHIC dataset does not include structured variables for leukocyte morphohlogy, so it was not possible to extract the criterion “immature [band] forms”. However, we assumed that in the majority of cases the leukocyte criterion would be met based on absolute numbers.
<sup>2</sup> We used vasopressor treatment rather than blood pressure for the definition of “hypotension”, because the criterion required that the patient had persistent hypotension despite adequate fluid resuscitation and required vasopressor treatment, and we assumed that the stan-

In Fig. 2, we present how the eligibility conditions were formally designed and how they can be specified based on the criteria description, from which each condition within the selection criteria is analysed regarding the aspects to be considered when specifying the corresponding logical conditions (see Table 1 for a full description of the selection criteria for the LeoPARDS trial).

**Figure 2:**
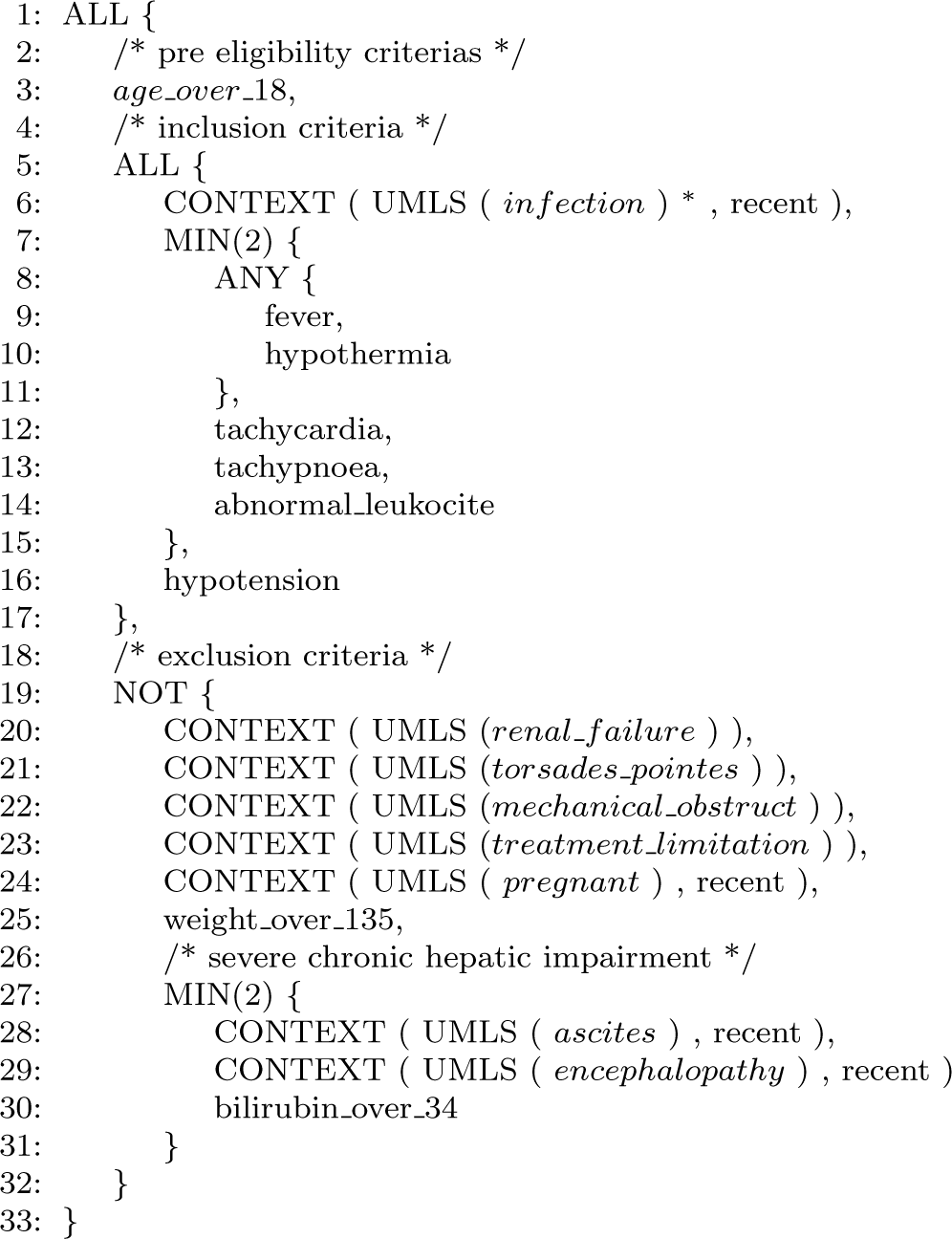
LeoPARDS selection criteria definition.^*^ See Table 4 for UMLS concepts defining *infection*

Table 4 lists the relevant UMLS concepts used to compound the definition of infection. The identifiers were collected from the existing UMLS concepts produced by the SemEHR annotation tool. Finally, Table 5 presents the UMLS concepts used to define other medical conditions described in the selection criteria for LeoPARDS.

**Table 3:** Comparison of demographic characteristics between patients recruited in the original LeoPARDS trial and those identified by the *CogStack* screening process.

| Demographic | LeoPARDS | LeoPARDS | CogStack |
| --- | --- | --- | --- |
| Attribute | Levosimendan | Placebo | Screening |
| Median age (IQR) yr | 67 (58–75) | 69 (58–77) | 66 (54–77) |
| Median weight (IQR) kg | 76 (65–90) | 80 (68–91) | 70 (60–82) |
| Male sex (%) | 56.2% | 56.0% | 59.7% |

**Table 4:**
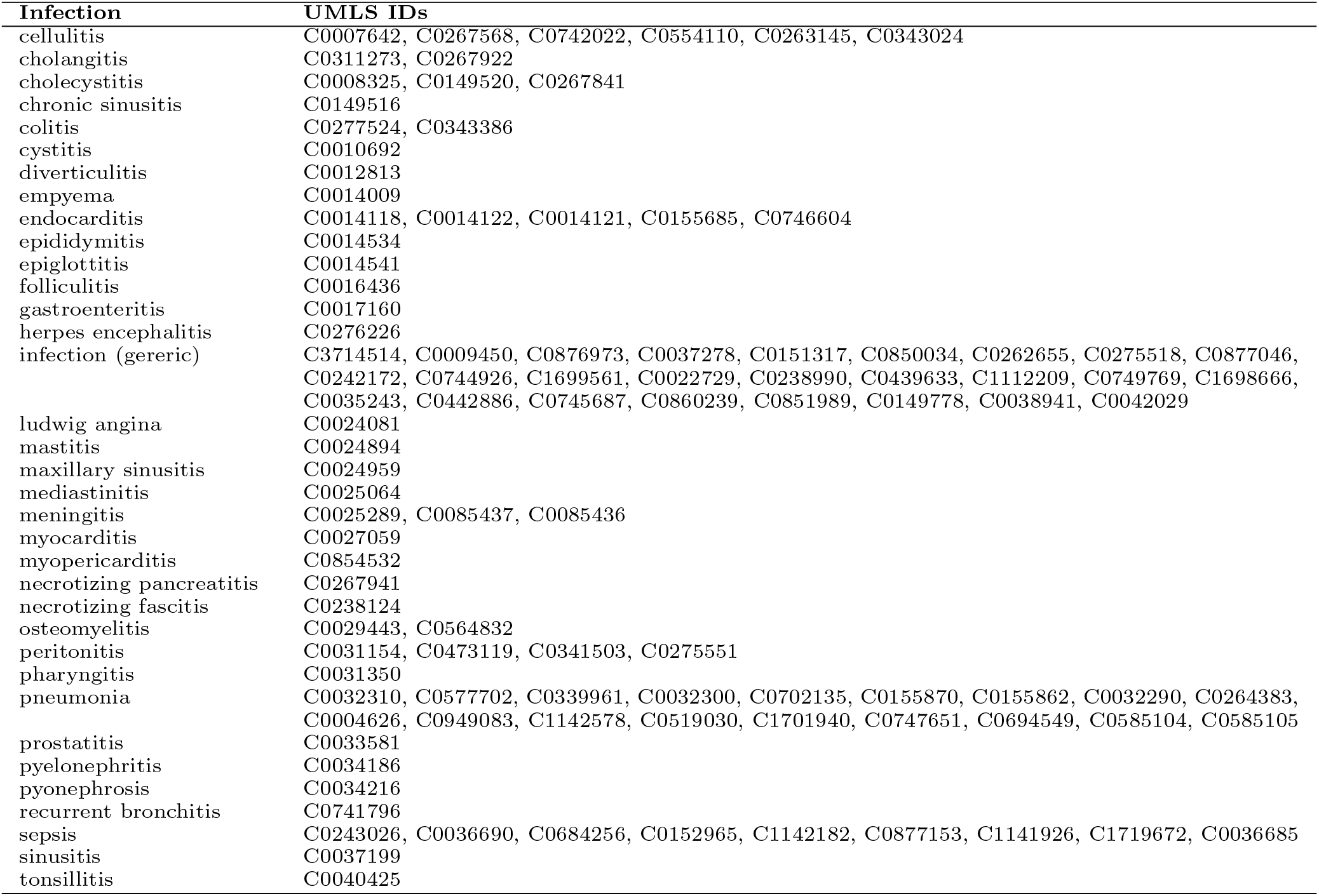
UMLS relevant concepts for “infection”.

| Infection | UMLS IDs |
| --- | --- |
| cellulitis | C0007642, C0267568, C0742022, C0554110, C0263145, C0343024 |
| cholangitis | C0311273, C0267922 |
| cholecystitis | C0008325, C0149520, C0267841 |
| chronic sinusitis | C0149516 |
| colitis | C0277524, C0343386 |
| cystitis | C0010692 |
| diverticulitis | C0012813 |
| empyema | C0014009 |
| endocarditis | C0014118, C0014122, C0014121, C0155685, C0746604 |
| epididymitis | C0014534 |
| epiglottitis | C0014541 |
| folliculitis | C0016436 |
| gastroenteritis | C0017160 |
| herpes encephalitis | C0276226 |
| infection (gereric) | C3714514, C0009450, C0876973, C0037278, C0151317, C0850034, C0262655, C0275518, C0877046, C0242172, C0744926, C1699561, C0022729, C0238990, C0439633, C1112209, C0749769, C1698666, C0035243, C0442886, C0745687, C0860239, C0851989, C0149778, C0038941, C0042029 |
| ludwig angina | C0024081 |
| mastitis | C0024894 |
| maxillary sinusitis | C0024959 |
| mediastinitis | C0025064 |
| meningitis | C0025289, C0085437, C0085436 |
| myocarditis | C0027059 |
| myopericarditis | C0854532 |
| necrotizing pancreatitis | C0267941 |
| necrotizing fascitis | C0238124 |
| osteomyelitis | C0029443, C0564832 |
| peritonitis | C0031154, C0473119, C0341503, C0275551 |
| pharyngitis | C0031350 |
| pneumonia | C0032310, C0577702, C0339961, C0032300, C0702135, C0155870, C0155862, C0032290, C0264383, C0004626, C0949083, C1142578, C0519030, C1701940, C0747651, C0694549, C0585104, C0585105 |
| prostatitis | C0033581 |
| pyelonephritis | C0034186 |
| pyonephrosis | C0034216 |
| recurrent bronchitis | C0741796 |
| sepsis | C0243026, C0036690, C0684256, C0152965, C1142182, C0877153, C1141926, C1719672, C0036685 |
| sinusitis | C0037199 |
| tonsillitis | C0040425 |

**Table 5:** Other UMLS concepts used to compound the LeoPARDS selection criteria.

| Concept | UMLS IDs |
| --- | --- |
| torsades_pointes | C0040479, C1960156, C1963250, C3150851, C4510938, C4510799, C4511461 |
| renal_failure | C0011946, C0015354, C0019004, C0019014, C0022661, C0031139, C0041612, C0191116, C0200017, C0206075, C0264654, C0268810, C0271932, C0398312, C0398338, C0398340, C0398343, C0398344, C0403462, C0403463, C0403464, C0403465, C0419061, C0419062, C0455667, C0558708, C0565539, C0748315, C1561829, C3494724, C3531744, C3536572, C3649547, C3697607, C4038741, C4047993 |
| mechanical_obstruct | C0003492, C0003499, C0003507, C0024164, C0026269, C0151241, C0152417, C0155567, C0158618, C0264766, C0264772, C0275846, C0332886, C0340335, C0340361, C0340371, C0340372, C0340373, C0340375, C0344401, C0345086, C0345087, C0349073, C0349075, C0349516, C0406810, C0700637, C1290389, C1306822, C1850635, C1868705, C1960800, C3532372, C3532376, C3839320, C3839383, C3839635 |
| treatment_limitation | C0582114, C3472262, C4305111 |
| liver_impairment | C0019147, C0019212, C0085605, C0162557, C0274386, C0400927, C0400928, C0400929, C0745744, C1619727, C2936476, C4039103 |
| pregnant | C0026751, C0032979, C0032980, C0032981, C0032995, C0033150, C0041747, C0149973, C0232989, C0232990, C0232991, C0232992, C0232993, C0232994, C0242786, C0269675, C0278056, C0404831, C0404842, C0425965, C0425979, C0425983, C0425984, C0425985, C0425986, C0425987, C0549206, C0585066, C0860096, C1291689, C2586154 |
| ascites | C0003962, C0008732, C0019086, C0025184, C0031144, C0220656, C0267772, C0267773, C0267774, C0267776, C0269720, C0275919, C0341525, C0401037, C0401038, C0437001, C0585187, C0741244, C1285291, C3532188, C3665480, C4038874, C4038944 |
| encephalopathy | C0019147, C0019151, C3266165 |

### 2.6 Comparison of Automated and Manual Screening

We compared the set of patients identified as eligible for LeoPARDS by the new algorithm with the screening logs for the original trial. For patients detected as eligible by the algorithm but not screened in the original trial, we carried out a manual case note review of a random sample. Two clinicians reviewed the original EHR case notes on the ICIP system to ascertain whether the algorithm correctly applied the eligibility criteria and what the likely clinical reason that the patient was not included in the screening log.

## Results

In the results presented in this section we used the terms *LeoPARDS* to indicate results from the original trial, *Screening* to refer to the original manual screening log from UCLH, *CogStack* to refer to the results found by our mimicking application, and *Overlapping* to designate those patients that were found in both the *Screening* and the *CogStack* results.

For the actual *LeoPARDS* trial in UCLH, there were 315 *Screening* episodes for 303 distinct patients (some were screened more than once), and 47 patients were recruited. Seven of these patients lacked structured data on the date of screening because of incomplete data extraction in CCHIC, leaving 40 for the comparison with *Cogstack*.

We used the concept of “episode” as the fundamental EHR entity search in *CogStack*, which comprises all the data being recorded during the ICU stay. Each episode also contains the demographic information of the patient, ward transferring origin and destination within a hospital and diagnosis information. *CogStack* was able to find 407 candidate episodes, corresponding to 395 distinct screening patients (we only considered the first episode from each patient matching the selection criteria). The *Overlapping* set between *CogStack* and UCLH *Screening* corresponds to 208 patients, of which 155 had a screening date which matched within one day between the manual *Screening* log and *CogStack*.

Fig. 3 shows the numbers of screened and recruited patients by month from June 2014 to December 2015. Of the 84 *Screening* episodes (83 patients) not detected by *Cogstack*, 60 had no CCHIC structured data available. CCHIC structured data were incomplete for the third quarter of 2015 because NHS audit activities were suspended between July and August 2015 due to staffing shortages. It was expected that the *CogStack* algorithm would not be able to identify eligible patients during this period. Among patient episodes that had CCHIC structured data available, the *Cogstack* algorithm detected all 40 who were recruited in the actual *LeoPARDS* trial.

**Figure 3:**
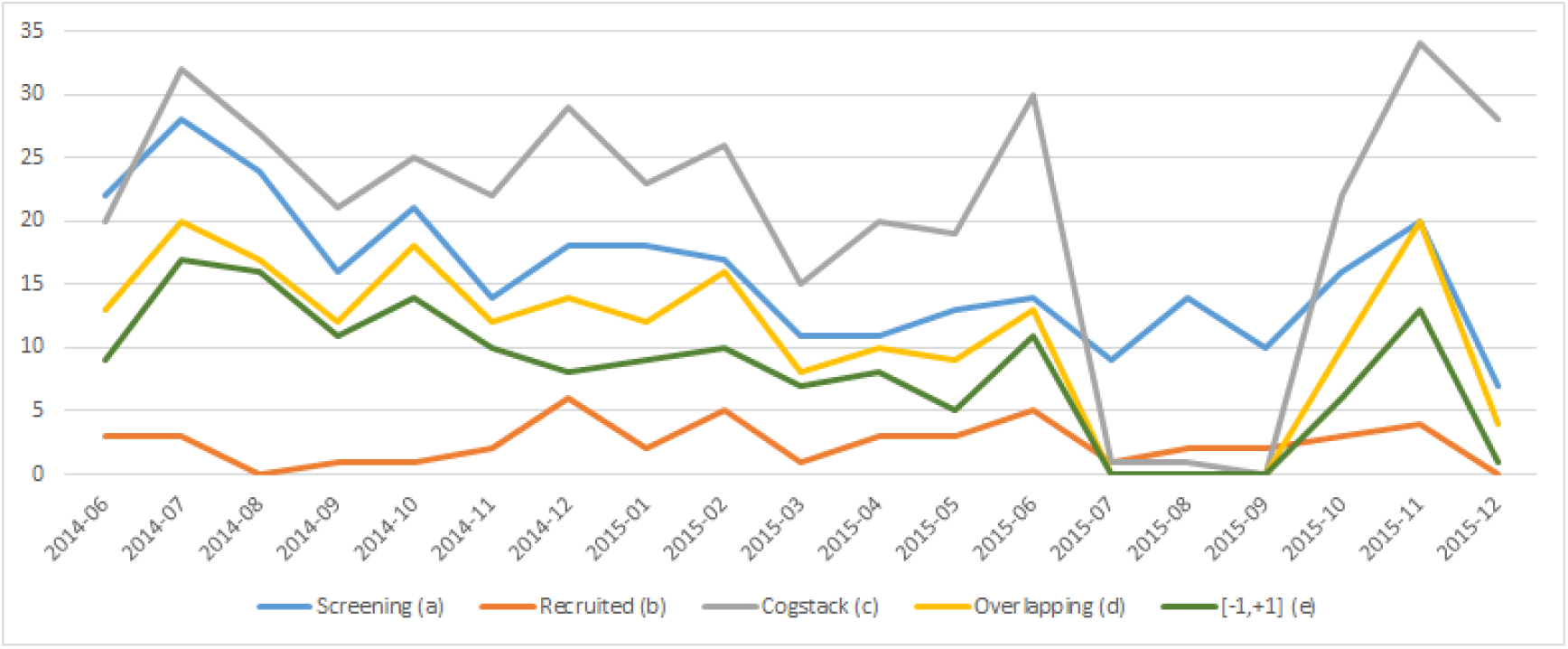
(a) number of patients in the original UCLH *Screening* log; (b) number of patients recruited in UCLH; (c) number of patients found by *Cogstack*^*^; (d) number of patients overlapping between *CogStack* and the original UCLH *Screening* log; (e) number of patients overlapping with up to 1 day difference between the screening date from UCLH *Screening* log and *CogStack* – ^*^ *CogStack* was not able to find patients matching the selection criteria between July and August 2015 due to a known lack of data in the CCHIC dataset.

The distribution of age and gender in patients identified by *CogStack* was consistent with patients recruited to the original LeoPARDS trial, as shown in Table 3 and Fig. 4.

**Figure 4:**
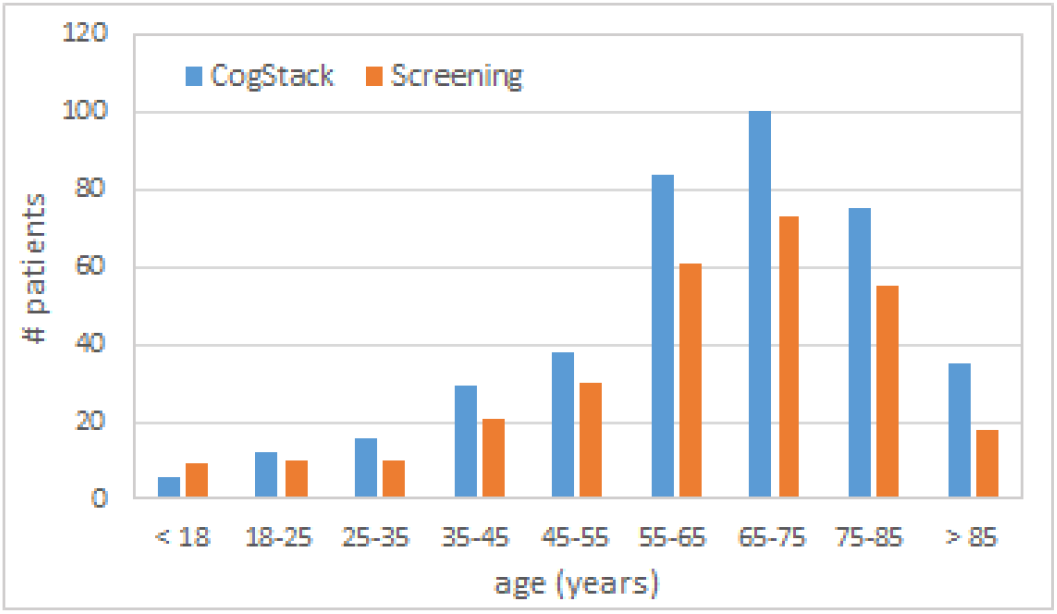
Age distribution of patients identified by *CogStack* as being eligible for the trial.

**Figure 5:**
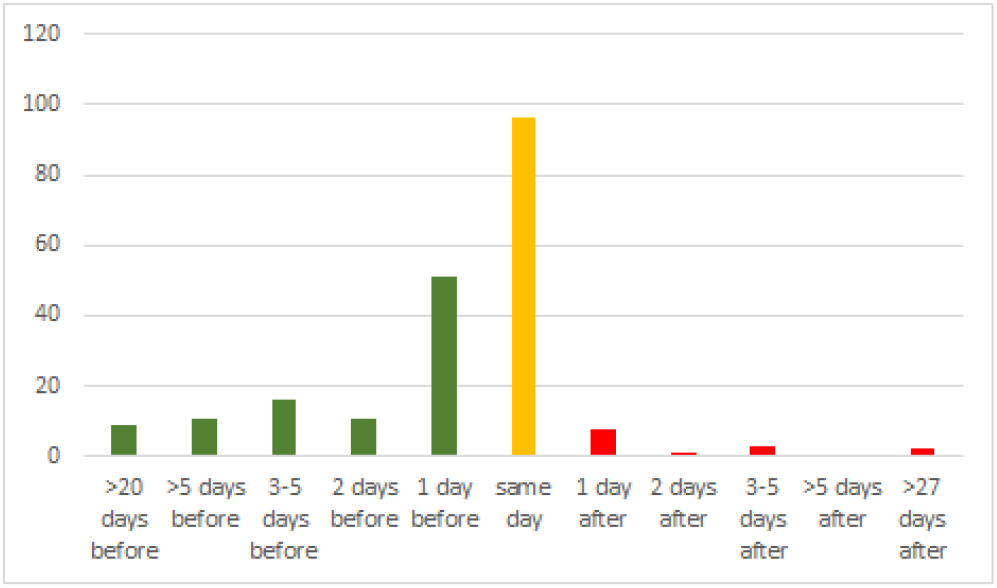
Timing of eligibility identified by *CogStack* compared to manual screening.

We also analysed the ability of *CogStack* to identify eligible patients earlier than the original UCLH *Screening* log. From the 208 *Overlapping* patients, 96 were found by *CogStack* the same day as the original screening, 62 patients were detected one or two days earlier, and 36 patients were detected three or more days earlier than the *Screening* log. Where *CogStack* was not able to identify patients as early as the manual screening log, this was because they had been matched to the same patient in an earlier or later critical care episodes.

Among patients detected by *Cogstack* but not screened in the original trial, we manually reviewed the clinical notes of a random sample of 16/147 (10.9%). We found only 2 patients (13%) who could potentially have been enrolled based on information gleaned from detailed review of the clinical notes, and one of these was not screened because it was during the New Year holiday period when trial staff were not working. Half of the patients (8/16) strictly met the inclusion and exclusion criteria, but were considered clinically unlikely to benefit from an experimental therapy because they were either too sick or dying (five patients) or at the least severe end of the spectrum (three patients). Inclusion of such patients in the trial would dilute the power of the trial, and would not be within the patient group that might be treated in the future if the trial showed a positive outcome. Six patients (38%) had an alternative explanation for the combination of antibiotic treatment and physiological parameters that suggested sepsis, but which were not programmed into the Cogstack algorithm. A typical example was a post-surgical patient on prophylactic antibiotics, with raised respiratory rate and heart rate (possibly due to pain), requiring inotropes for hypotension due to an epidural.

### Performance

Finally, in terms of processing time, *CogStack* demonstrated potential to serve as a near real-time search and filtering tool in order to facilitate the pre-screening process. The full process of screening 11,500 24-hour-sliding windows (during the 16 months from which substantial data was available) was performed in less then 15 minutes (890 seconds) – corresponding to less than 0.1 second per window screening. Experiments were performed using a Linux server Intel® Xeon® 8 CPUs 64-bits E5-2680 v4 2.40GHz 64GB RAM.

## Discussion

This study showed that an electronic algorithm incorporating NLP could successfully match patients against the selection criteria for a clinical trial in the critical care setting with a time-sensitive recruitment window. It suggests that such methods may potentially be useful for automatic pre-screening of potential clinical trial participants, reducing the amount of manual input required for this process. Although taking into account only a subset of free text notes, CogStack was able to find a considerable overlapping along the set of patients originally included in the UCLH screening log, and the patients found by CogStack followed the demographic distribution reported in the LeoPARDS trial. Besides being faster, CogStack was also able to identify almost 10% of the overlapping patients that were included in the automatic screening report in the range of 1-3 days earlier comparing to manual screening, from which the eligibility explicitly states the 24-hour windows for recruitment was missed.

Results from the manual check showed that strict application of the criteria resulted in some patients being identified who would not be included based on clinical judgement (if they were not sick enough to risk an experimental treatment, or if they were so sick that any intervention was likely to be futile). This suggests that trial inclusion and exclusion criteria need to be more explicit if they are to be accurately applied by computer algorithms while truly capturing the desired patient population. Very few additional eligible patients were detected by the CogStack algorithm, which shows that the manual processes for participant identification were thorough, albeit resource-intensive.

### Improving efficiency of clinical trials

By virtue of randomisation, randomised controlled trials (RCTs) are considered the gold standard to assess the effects of medical interventions such as pharmacological treatments [19]. However, RCTs are also notorious for their time-consuming nature, high costs, and the fact that the populations included in RCTs often do not resemble real-world patient populations [1, 5, 7]. Registry-based trials have been proposed to overcome some of these limitations, while maintaining the scientific rigour of randomisation [20–22]. In registry-based trials, existing EHR data registries (electronic health record databases) are used for patient recruitment and follow-up, while the experimental intervention is randomly assigned, as in a conventional RCT [20, 21]. Since these studies are executed in routine clinical practice, their results tend to better reflect effectiveness in clinical practice [20, 23]. Another advantage of EHR-based automatic patient selection is that the algorithms can be modified and re-applied to test different patient selection criteria, making it easier to design future trials [24].

Clinical trials need to recruit participants according to the eligibility criteria defined in the study protocol in order to accurately answer the question they set out to. Trial sites usually spend most of their time on patient recruitment, and yet, statistics show that, despite their efforts, reaching enrolment goals per timelines seems elusive in many studies, with over 80% of clinical trials failing to meet enrolment timelines [25]. Among randomised controlled trials funded by the NIHR Health Technology Assessment programme, the final recruitment target sample size was achieved in only 56% [26]. This can have major impact on the feasibility, power and validity of the trials.

There are a number of challenges in using electronic health records for identifying trial participants. Mapping the selection criteria to logical conditions can be difficult, as eligibility criteria are described using natural language designed for human rather than computer interpretation, and need to be translated into complex queries running on multiple EHR data sources. Representation of time constraints also needs to be taken into account [27]. Temporal references can be described in diverse ways with varying degrees of precision (e.g. “within the previous 24 hours”, “previous treatment within 30 days”, “for at least four hours”) [28].

### Natural language processing

Detailed information on patient characteristics that are relevant to trial inclusion and exclusion criteria may be present only in the free text of EHRs. Although narrative data is a valuable asset for improving healthcare, it is usually inaccessible due to its lack of structure, hence the need for natural language processing (NLP) applications to extract information in a structured form. A diverse set of NLP applications exist in the clinical domain including: (i) identifying complications among intensive care unit patients [29], (ii) collecting uniform data in routine clinical practice for optimal care, quality control and research [30], (iii) using machine learning approaches for clinical notes classification [31], and (iv) increasing the efficiency of automated clinical trial eligibility [8].

Despite recent progress in more sophisticated NLP tools, extracting data from clinical notes remains challenging. A systematic review [32] presents existing NLP systems that generate structured information from unstructured clinical free text, describing 86 papers fitting the review criteria and containing information about 71 different clinical NLP systems. Most of the approaches to date have a fairly narrow focus using simple rule-based approaches (e.g. regular expression patterns) in order to address specific information extraction tasks, but they require extensive human intervention for application to new tasks. Machine learning approaches have been growing in popularity, aided by the increasing number of publicly available clinical datasets for training algorithms.

Text analytics platforms such as semEHR (built on CogStack) [13, 14] and GATE [33] are increasingly being used across large document repositories, and can incorporate a range of NLP methods such as Bio-Yodie [15] (rules-based information extraction, used in this project) and machine learning metehods. UCLH is proposing to make semEHR a core component of its new clinical research data warehouse. UCLH is building infrastructure for handling unstructured data, following the examples of King’s College Hospital (KCH) and the South London and Maudsley (SLAM) mental health Trust.

### Strengths and limitations

The main strength of this study was the demonstration of algorithms combining structured EHR data and NLP to assist participant recruitment in a simulation of a real clinical trial. The LeoPARDS trial had particular recruitment challenges – the time-sensitive nature of the task, and the severity of the patients’ condition.

A limitation was that our algorithm attempted to identify a diagnosis of sepsis which may be difficult even for experienced clinicians. Hence application of the strict inclusion and exclusion criteria identified patients who were not eligible because they had an alternative explanation for their physiological state that was not sepsis; this was apparent to clinician reviewers but not to the algorithm because it was not programmed in. This highlighted the need for much more explicit trial inclusion algorithms if they are to be interpreted automatically, and it may be difficult to plan for all such nuances in advance.

Our algorithm was limited in that it only included key portions of the free text rather than the entire clinical record, and the identification of some criteria was not possible (such as white cell morphology). We were limited to a single site because only the UCLH site currently had free text available for NLP, but the method could potentially be scaled to many sites and adapted for different studies.

### Clinical and research implications

This study has demonstrated the feasibility of this approach in a critical care trial. Future work should apply this method at other sites and for other studies, and to develop a method for a current clinical trial in order to evaluate its utility and performance for real-time patient screening and recruitment.

The algorithm could also be tuned by testing out different thresholds for inclusion and exclusion, in order to achieve a combination of sensitivity and specificity which best suits its use in combination with manual review in a trial recruitment scenario. Use of EHR data with NLP could also be used to extract participant data for the trial case report forms. This will save even more time by avoiding the need for duplicate data entry, and enable the use of more detailed measures of health status, such as continuous monitoring of physiological parameters rather than a single measurement in a case report form. However, it also introduces new challenges such as ensuring validity, completeness and accuracy of the data [34, 35], and harmonising heterogenous data across institutes.

## Conclusions

Electronic health record data may potentially be used in computer algorithms to help identify trial participants and increase recruitment in clinical trials, but much of the detailed clinical information is available only in the form of free text. We simulated screening and recruitment for the LeoPARDS trial in critical care, by the Cogstack platform with natural language processing tools to process electronic health record data. Cogstack was able to identify the majority of patients originally screened, including all those recruited, and in many cases identify patients as eligible one or two days before the actual manual screening process. This approach could be implemented in real time to facilitate clinical trial recruitment, and reduce the burden of time-consuming manual case note review.

## Data Availability

Patient data analysed for this project was extracted from the Critical Care Health Informatics Collaboration research database, which has had National Research Ethics Service approval (14/LO/1031). Individual participant consent was not required, as section 251 exemption was granted by the Confidentiality Advisory Group of the Health Research Authority.

## Acknowledgements

This study was supported by the National Institute for Health Research (NIHR) University College London Hospitals (UCLH) Biomedical Research Centre (BRC) Clinical and Research Informatics Unit (CRIU), NIHR Health Informatics Collaborative (HIC), and by awards establishing the Institute of Health Informatics at University College London (UCL).

This study was funded Health Data Research UK (grant No. LOND1), which is funded by the UK Medical Research Council, Engineering and Physical Sciences Research Council, Economic and Social Research Council, Department of Health and Social Care (England), Chief Scientist Office of the Scottish Government Health and Social Care Directorates, Health and Social Care Research and Development Division (Welsh Government), Public Health Agency (Northern Ireland), British Heart Foundation and Wellcome Trust. ADS is supported by a postdoctoral fellowship from THIS Institute.

RD is supported by: (a) the National Institute for Health Research (NIHR) Biomedical Research Centre at South London and Maudsley NHS Foundation Trust and Kings College London. The views expressed are those of the author(s) and not necessarily those of the NHS, the NIHR or the Department of Health; (b) Health Data Research UK, which is funded by the UK Medical Research Council, Engineering and Physical Sciences Research Council, Economic and Social Research Council, Department of Health and Social Care (England), Chief Scientist Office of the Scottish Government Health and Social Care Directorates, Health and Social Care Research and Development Division (Welsh Government), Public Health Agency (Northern Ireland), British Heart Foundation and Wellcome Trust; (c) The BigData@Heart Consortium, funded by the Innovative Medicines Initiative-2 Joint Undertaking under grant agreement No. 116074. This Joint Undertaking receives support from the European Unions Horizon 2020 research and innovation programme and EFPIA; it is chaired, by DE Grobbee and SD Anker, partnering with 20 academic and industry partners and ESC; (d) The National Institute for Health Research University College London Hospitals Biomedical Research Centre.

FA is supported by UCL Hospitals NIHR Biomedical Research Centre.

DB is partially funded by the Division of Critical Care, University College Hospital and NIHR University College London Hospitals Biomedical Research Centre.

## Short Bios

**Hegler C. Tissot** is a Senior Research Fellow in Health Informatics. He received his Ph.D. degree in Computer Science from the Universidade Federal do Paran in 2016, as a member of the C3SL Labs, and was a postdoctoral researcher at Macquarie University (Australia), in 2017. He works with knowledge engineering and his main research interest is about improving public health services by applying natural language processing to extract structured information from electronic health records and using machine learning to design prediction models within the clinical domain.

**Anoop D. Shah** is a Clinical Lecturer at the UCL Institute of Health Informatics and Consultant in Clinical Pharmacology and General Medicine at UCLH. His PhD focused on blood biomarkers and cardiovascular diseases in linked electronic health record databases. He is a Fellow of the Faculty of Clinical Informatics, leading on work to improve diagnosis recording, and currently holds a post-doctoral research fellowship with The Healthcare Improvement Studies Institute.

**Ruth Agbakoba** is a Honorary Research Associate at the UCL Institute of Health Informatics and a Project Engagement Lead within the UCLH Clinical Research Informatics Unit. She is leading on the design, implementation and evaluation of an innovative clinical trials discovery platform to be deployed across the entire trust of UCLH hospitals. She has background in health informatics, digital health and eHealth, with current interests focused in translational research (translating research findings into practice) and Global eHealth policy to foster person-centred care.

**Amos Folarin** is the Senior Software Development Group Leader at the KCL BRC-MH. His background is in biochemistry/molecular biology. He is currently working on developing the RADAR-CNS data collection platform for remote patient monitoring using wearable devices, mobile phone sensors and mobile apps. His interests include monitoring seasonal infectious diseases, deep-learning image analysis pipelines for highcontent screening, and building a portable next generation sequencing pipeline for Genomics England.

**Luis Romao** is a Programme Manager for Clinical Research Informatics at the National Institute of Health Research (NIHR) Biomedical Research Centre (BRC) at UCLH/UCL and an Honorary Research Associate at the UCL Institute of Health Informatics. He currently manages the BRC UCLH/UCL Clinical Research Informatics Unit, including the NIHR Health Informatics Collaborative Programme at UCLH. He holds an MSc in Health Management in Strategic Management and Leadership from City University London. He was responsible for overseeing the implementation of EHR (EPIC) Systems across a number of international institutions, including Mount Sinai Hospital, New York, USA.

**David Brealey** is a Consultant in Anaesthesia and Intensive Care Medicine at University College London Hospitals NHS Foundation Trust. He is the Trust lead for Critical Care, Anaesthesia and Emergency Medicine Research. His earlier research was the first to demonstrate mitochondrial dysfunction as a potential cause of sepsis induced organ failure in patients. He now leads a successful Critical Care clinical trials team which are recognised as one of the highest performing teams within the UK and capable of performing on a global level.

**Steve Harris** is a Critical Care physician. His clinical training included anaesthesia and tropical medicine, and he worked for Medecins Sans Frontieres in Congo-Brazzaville, the Democratic Republic of Congo and Haiti. In 2009, he won a Wellcome Clinical Research Training Fellowship. He was awarded my PhD from LSHTM in 2014, and became an National Institute of Health Research (NIHR) Clinical Lecturer at University College London (UCL) before being appointed as a Consultant in Critical Care at University College Hospital London (UCLH) in 2016. He now leads the software development for the NIHR Health Informatics Collaborative Critical Care theme.

**Lukasz Roguski** is as Software Developer in charge of the CogStack project at University College London (UCL), UK, an information retrieval and extraction platform for unlocking electronic health records. Prior to joining UCL, he worked in the National Center for Genomic Analysis (CNAG-CRG) in Barcelona, Spain, where he has been developing methods for high-throughput sequencing data compression. He has graduated with PhD in Bioinformatics from Universitat Pompeu Fabra (UPF) in Barcelona, Spain.

**Richard Dobson** is Professor of Medical Informatics and Head of Bioinformatics at the NIHR Biomedical Research Centre for Mental Health (KCL) and the South London and Maudsley NHS Trust. His main areas of bioinformatics research have focused on the genomics of complex disease, with a special focus on biomarkers of Alzheimer’s Disease. Research has required the analysis, integration and modeling of complex large molecular datasets.

**Folkert W. Asselbergs** is Prof in cardiovascular genetics and consultant cardiologist at the department of Cardiology, University Medical Center Utrecht, Prof of Precision medicine at the Institute of Cardiovascular Science and Institute of Health Informatics, University College London, Director BRC Clinical Research Informatics Unit, University College London Hospital, Manager Research Center for Circulatory Health, UMC Utrecht, and chair data infrastructure Dutch Cardiovascular Alliance (www.dcvalliane.nl). Lately, he widened his research focus to precision medicine using linked data sources such as wearable information and routine care data obtained from electronic health records including free text. His ambition is to build a network for performing clinical trials within routine health care linked to national registries.

## Biographical Note

The Institute of Health Informatics (IHI) is an academic department at UCL within the Faculty of Population Health Sciences (FPHS), at the School of Life & Medical Sciences (SLMS). IHI works in partnership with the UCLH Biomedical Research Centre (BRC), funded by the National Institute for Health Research, aiming to support translational and health service research.

## Key Points

- Identification of suitable participants for clinical trials is a resource-intensive process, and particularly difficult for time-critical trials, usually requiring manual screening of large numbers of clinical notes.
- Automated methods to identify suitable patients using electronic health records are limited by the lack of structured information in the records.
- We found that natural language processing of unstructured data, combined with algorithms applied to structured data, could successfully simulate the screening and recruitment process for the LeoPARDS trial of a treatment for sepsis.

## Footnotes

1 http://www.elastic.co/

2 http://www.timeml.org/

